# Altered pre-existing SARS-CoV-2-specific T cell responses in elderly individuals

**DOI:** 10.1101/2021.07.26.21261082

**Authors:** Naoyuki Taira, Sakura Toguchi, Mio Miyagi, Tomoari Mori, Hiroaki Tomori, Koichi Oshiro, Osamu Tamai, Mitsuo Kina, Masatake Miyagi, Kentaro Tamaki, Mary K Collins, Hiroki Ishikawa

**Author notes:** Corresponding author Hiroki Ishikawa PhD.

## Abstract

Pre-existing SARS-CoV-2-specific T cells, but not antibodies, have been detected in some unexposed individuals. This may account for some of the diversity in clinical outcomes ranging from asymptomatic infection to severe COVID-19. Although age is a risk factor for COVID-19, how age affects SARS-CoV-2-specific T cell responses remains unknown. We found that some pre-existing T cell responses to specific SARS-CoV-2 proteins, Spike (S) and Nucleoprotein (N), were significantly lower in elderly donors (>70 years old) who were seronegative for S than in young donors. However, substantial pre-existing T cell responses to the viral membrane (M) protein were detected in some elderly donors. These responses likely compensate for loss of T cell responses specific to S and N. In contrast, young and elderly donors exhibited comparable T cell responses to S, N, and M proteins after infection with SARS-CoV-2. M-specific responses were mediated by CD4 T cells producing interferon-γ in both seronegative and seropositive individuals. T cells in seronegative elderly donors responded to various M-derived peptides, while the response after SARS-CoV-2 infection was apparently focused on a single peptide. These data suggest that diversity of target antigen repertoire for pre-existing SARS-CoV-2-specific T cells declines with age, but the magnitude of pre-existing T cell responses is maintained by T cells reactive to specific viral proteins such as M. A better understanding of the role of pre-existing SARS-CoV-2-specific T cells that are less susceptible to age-related loss may contribute to development of more effective vaccines for elderly people.

## Introduction

There is extensive individual variation in severity of coronavirus disease 2019 (COVID-19), caused by severe acute respiratory syndrome coronavirus 2 (SARS-CoV-2), ranging from asymptomatic infection to fatal pneumonia (1). Various factors, including age, sex, and comorbidities such as obesity and diabetes, influence the risk of severe COVID-19 (2–4). For example, morbidity and mortality among the elderly are significantly higher than among the young (2). Consideration of protective measures for individuals vulnerable to COVID-19 should be particularly important to control the pandemic (5). However, the cellular and molecular bases of the variable risk of COVID-19 remain poorly understood.

T cells are assumed to mediate both protective and pathogenic immune responses to SARS-CoV-2 infection (6, 7). The magnitude and quality of T cell responses induced by SARS-CoV-2 infection are highly heterogeneous and are likely associated with COVID-19 clinical outcomes. For example, SARS-CoV-2-specific T cell numbers and their interferon-γ (IFN-γ) expression in severe COVID-19 patients are lower than in mild COVID-19 patients (8–10). Furthermore, asymptomatic COVID-19 patients tend to have increased SARS-CoV-2-specific T cells expressing higher levels of IFN-γ compared to symptomatic patients (11). This individual variation in T cell responses may be partly explained by heterogeneity in levels of pre-existing SARS-CoV-2-reactive T cells.

Some individuals who have not been exposed to SARS-CoV-2 have nonetheless acquired SARS-CoV-2-reactive T cells, probably through exposure to other common cold coronaviruses (12–14). Pre-existing CD4 and CD8 memory T cells, specific to various SARS-CoV-2 proteins, including the structural proteins, Spike (S), Membrane (M), and Nucleoprotein (N), have been detected with significant individual variation (D. Wyllie et al., manuscript posted on medRxiv DOI: 10.1101/2020.11.02.20222778). These pre-existing SARS-CoV-2-reacive T cells are associated with immune protection against COVID-19; however, in other cases, they may exacerbate COVID-19 severity (15, 16). As many of the current vaccines express the SARS-CoV-2 S protein, only pre-existing S-reactive T cells are activated by these vaccines (Refs. 17 and L. Loyal et al., manuscript posted on medRxiv DOI: 10.1101/2021.04.01.21252379). Several studies have reported age-related differences in SARS-CoV-2-specific T cell responses in COVID-19 patients (Refs. 9 and C. A. Cohen et al., manuscript posted on medRxiv DOI: 10.1101/2021.02.02.21250988); however, the effect of age on pre-existing SARS-CoV-2-reactive T cells remains unknown.

In this study, we compared frequencies of T cells reactive to SARS-CoV-2 S, N, and M antigens between young and elderly donors. The relatively elderly Okinawan population, and the moderate rate of SARS-CoV-2 infection in Okinawa, allowed us to examine pre-existing SARS-CoV-2-specific T cells in a cohort of elderly (>70 years old) individuals. We found that pre-existing T cell responses to S and N antigens are significantly impaired in elderly donors compared to young donors, but a proportion of elderly donors exhibit significant, high levels of M-reactive T cell responses. These data provide new insights into age-related alteration of pre-existing SARS-CoV-2-specific T cells.

## Methods

### Subjects

The study design was approved by the Okinawa Institute of Science and Technology, Graduate University (OIST) human subjects ethics committee (applications HSR-2020-024, HSR-2020-028). All donors provided informed written consent. Young (20 to 50 years of age, n=66) and elderly volunteers (over 70 years of age, n=52) were recruited in Okinawa, Japan, between October, 2020 and April, 2021. 90 donors (48 young and 42 elderly) had no history of COVID-19, while 28 donors (18 young and 10 elderly) who recovered from COVID-19 had positive COVID-19 PCR test results 1-3 months before blood collection. Plasma from each donor was tested for SARS-CoV-2-specific antibodies using SARS-Cov-2 Antibody Detection Kits (KURABO RF-NC001, RF-NC002) or Cellex qSARS-Cov-2 IgG/IgM Cassette Rapid Tests (Cellex 5513C). Four (3 young and 1 elderly) of 90 donors who had no history of COVID-19 and 26 (16 young and 10 elderly) of 28 donors who had recovered from COVID-19 were seropositive for SARS-CoV-2 S antigen. Based on antibody test results, donors were grouped into seronegative young (n=45, 40 % male, 60% female; mean age 38 years, age rage 23-49 years), seronegative elderly (n=41, 17 % male, 83% female; mean age 81 years, age rage 70-93 years), seropositive young (n=19, 63 % male, 37% female; mean age 41 years, age rage 20-50 years), and seropositive elderly (n=11, 45 % male, 55% female; mean age 78 years, age rage 70-91 years).

### Peripheral blood mononuclear cells (PBMCs) and plasma isolation

Blood samples were collected in heparin-coated tubes (TERUMO; VP-H100K). PBMCs and plasma were separated using Leucosep tupes pre-filled with Ficoll-Paque Plus (Greiner; 163288). After adding 5 mL of blood and 3 mL of AIM-V medium (Thermo; 12055091), Leucosep tubes were centrifuged at 1,000 g at room temperature for 10 min. The white layer containing PBMCs was collected, washed with 10 mL AIM-V medium and centrifuged for 7 min at 600 g, followed by a second washing with centrifugation for 7 min at 400 g. PBMC pellets were resuspended in 500 μL CTL test medium (Cellular Technology Limited (CTL); CTLT-010). Fresh PBMCs were used for IFN-γ ELISpot assays. PBMCs used for flow cytometry analysis and epitope mapping analysis were stored with CTL-cryo ABC media (CTL; CTLC-ABC) in liquid nitrogen.

### IFN-γ ELISpot assay

Peptide pools for SARS-CoV-2 S (JPT; PM-WCPV-S-1), N (Miltenyi;130-126-698), and M (Miltenyi;130-126-702) proteins dissolved in DMSO (500 μg/mL for S) or water (50 μg/mL for N and M) were used for cell stimulation. IFN-γ ELISpot assays were performed using Human IFN-γ Single-Color Enzymatic ELISpot kits (CTL; hIFNgp-2M), according to the manufacturer’s instructions. Briefly, freshly isolated PBMCs (1-4 x 10^5^ cells per well) were stimulated with 1μg/mL peptide solutions for each SARS-CoV-2 protein for 18-20 h. For each sample analysis, negative controls (cells treated with equimolar amounts of DMSO) and positive controls (cells treated with 50 ng/mL phorbol 12-myristate 13-acetate (PMA) and 1 μg/mL ionomycin) were included. After incubation, plates were washed and developed with detection reagents included in the kits. Spots were counted using a CTL ImmunoSpot S6 Analyzer. Antigen-specific spot counts were determined by subtracting background spot counts in a negative control well from the wells treated with peptide pools. If >30 spots/10^6^ PBMCs in the negative control well or <30 spots/10^6^ PBMCs in the positive control well were detected, sample data were excluded from analysis.

### Flow cytometry

Frozen PBMCs were thawed, washed with CTL wash supplement (CTL; CTL-W-010), and rested in CTL test medium overnight. Then, cells were resuspended in RPMI1640 (Gibco) medium supplemented with 5% (v/v) human AB-serum (PAN-Biotech; P30-2901), seeded into 96-well, U-bottom culture plates (10^6^ cells per well), and either left unstimulated (cells treated with equimolar amounts of DMSO) or stimulated with 1μg/mL SARS-CoV-2 M peptide pool for 7 h in the presence of 1μg/mL anti-CD40 (5C3; Biolegend; 334302) and 1 μg/mL anti-CD28 antibodies (CD28.2; Biolegend; 302934). Brefeldin A (1μg/mL) (Biolegend; 420601) was added for the last 2 h. After stimulation, cells were incubated with anti-Fc receptor-blocking antibody (Biolegend; 422301) and NIR-Zomibie (Biolegend; 423106) and stained with anti-CD3 (OKT3; Biolegend; 1:200), anti-CD4 (clone PPA-T4; Biolegend; 1:200), anti-CD8 (SK1; Biolegend; 1:200), anti-CD45RA (HI100; Biolegend; 1:100), and anti-CCR7 (G043H7; Biolegend; 1:100) antibodies. For intracellular cytokine analysis, cells were subsequently fixed and permeabilized using Foxp3 Staining Buffer Sets (eBioscience; 00-5253-00) and stained with anti-IFN-γ (B27; BD; 1:100), anti-TNF-α (MAb11; Biolegend; 1:20) and anti-IL-2 (MQ1-17H12; Biolegend; 1:20) antibodies. Samples were analyzed on a Fortessa X-20 (BD), and data were analyzed with FlowJo software version 10.7.1 (FlowJo LLC).

### Matrix peptide pools of SARS-CoV-2 M

Matrix peptide pools included in Epitope Mapping Peptide Set SARS-CoV-2 (VME1) (JPT EMPS-WCPV-VME-1) were used to analyze M epitopes recognized by T cells. PBMCs (0.5-1.5 x 10^5^) were stimulated with 1 μg/mL of each M matrix peptide pool (15 pools of 6-8 peptides) for 18 h and subjected to IFN-γ ELISpot assays.

### Statistical analysis

Unpaired t tests or Mann-Whitney U tests were performed using GraphPad Prism 9.1.0 software. Statistical details are provided in figure legends.

## Results

### Age-related differences in SARS-CoV-2-specific T cell responses

To assess whether there are age-related differences in SARS-CoV-2-specific T cell responses, we collected peripheral blood from young (20 to 50 years of age) and elderly (>70 years of age) donors in Okinawa between October 2020 and April 2021. These included 18 young and 10 elderly donors who had recovered from mild COVID-19 1-3 months prior to blood collection. Antibody tests using freshly purified sera showed that 93% of donors with previously diagnosed COVID-19 and 4% of those without, were seropositive for SARS-CoV-2 Spike (S). Then we divided donors into 4 groups: young seronegative (n=45), elderly seronegative (n=41), young seropositive (n=19), and elderly seropositive (n=11).

To compare SARS-CoV-2-specific T cell responses between age groups we performed Interferon-γ (IFN-γ) ELISpot assays using freshly purified peripheral blood mononuclear cells (PBMCs) stimulated with each of 4 peptide pools covering the major viral structural proteins [N-terminal S (S1), C-terminal S (S2), Membrane (M), or Nucleoprotein (N)]. First, we analyzed the sum of spots formed by IFN-γ-expressing cells reactive to S1, S2, N, or M antigens (hereafter referred to as spots of SNM-reactive T cells), indicating the magnitude of SARS-CoV-2-specific T cell responses in each donor. Almost all seropositive donors exhibited strong T cell responses to SARS-CoV-2 antigens regardless of age; the frequency of SNM-reactive spots was >40 per 10^6^ PBMCs in 92% of seropositive donors. The frequency of SNM-reactive T cells in seronegative donors was more variable than in seropositive donors, but a substantial proportion of seronegative donors exhibit T cell responses comparable to those of seropositive donors (53% of young donors and 42% of elderly donors had >40 spots per 10^6^ PBMCs). In both seronegative and seropositive populations, there were no significant differences in the frequency of SNM-reactive T cells between young (seronegative; median:53, IQR:21-96, seropositive; meidan:312, IQR:75-1509) and elderly (seronegative; median:32, IQR:7-331, seropositive; meidan:778, IQR:351-2392) (Fig. 1A, 1B).

**Fig. 1.**
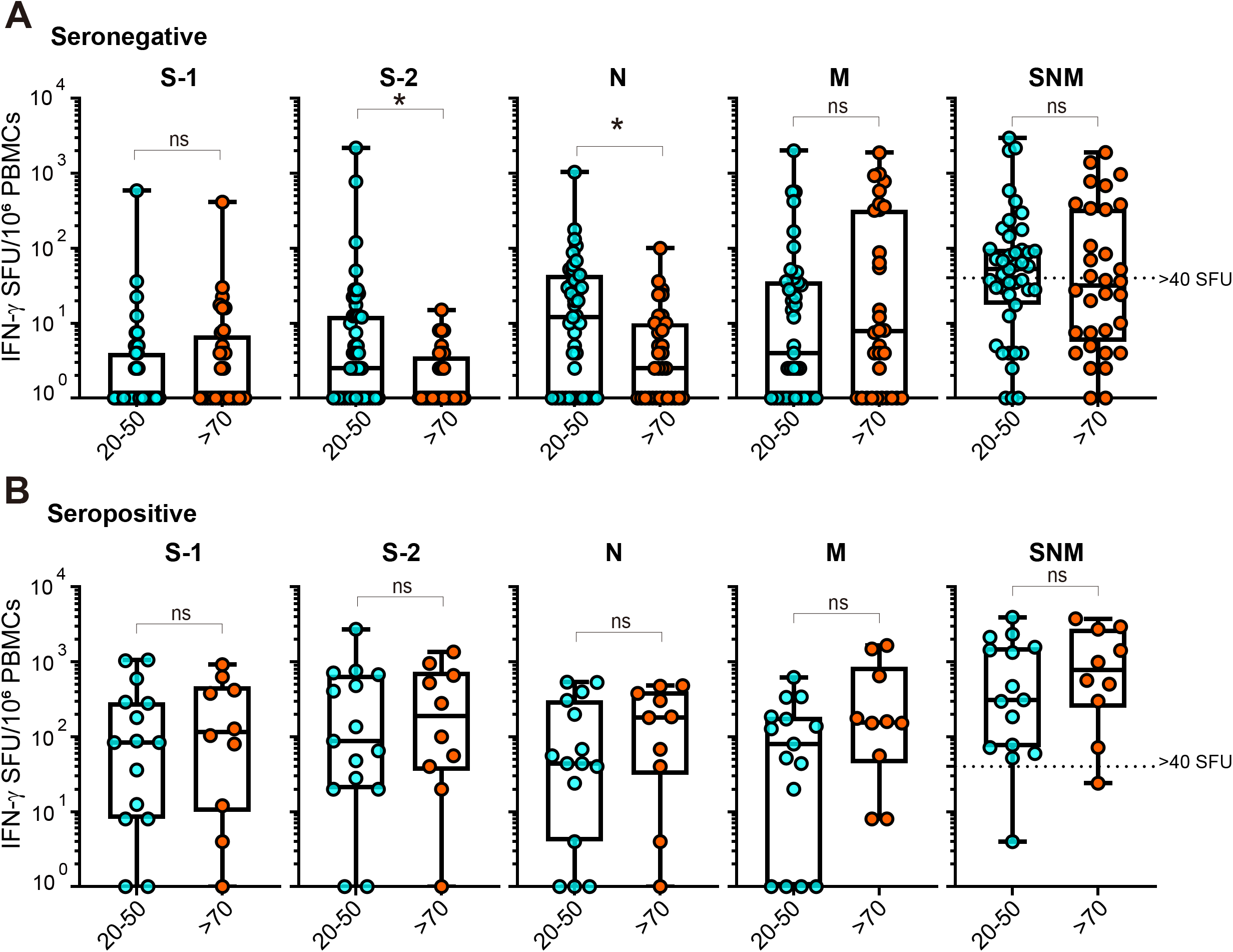
Altered pre-existing T cell responses to SARS-CoV-2 structural proteins in elderly donors. PBMCs isolated from seronegative (**A**) and seropositive (**B**) young (20-50 years of age) and elderly (>70 years of age) donors were stimulated with peptide pools for SARS-CoV-2 S, N, and M proteins and subjected to IFN-γ ELISpot analysis. Spot-forming units representing the frequency of IFN-γ-secreting cells in seronegative young (n=45), seronegative elderly (n=41), seropositive young (n=19), and seropositive elderly (n=11) are shown. The sum of spots formed by cells stimulated with S, N, and M (SNM) is also shown. Statistical comparisons between age groups utilized the Mann-Whitney test. *P<0.05, ns: not significant.

Next, we compared frequencies of T cells reactive to individual viral antigens between young and elderly donors. Among seronegative donors, the frequencies of S-2- and N-reactive T cells were significantly lower in elderly than in young persons (Fig. 1B). However, there were no significant differences in the frequencies of S-1- and M-reactive T cells between seronegative young and elderly donors (Fig. 1B). Consistent with previous reports (12), T cell responses to S-2 were higher than S-1 in the seronegative population (Fig. 1B). Frequencies of T cells specific to S1, S2, N, and M were comparable between seropositive young and elderly donors (Fig. 1B).

We also analyzed intraindividual immunodominance of each SARS-CoV-2 antigen in donors who had >40 spots per 10^6^ PBMCs. Remarkably, M-specific responses were dominant in 6/23 young and 12/14 elderly seronegative donors (Fig. 2A). In contrast, T cell responses specific to S-1 and S-2 were more prominent than M in seropositive donors (Fig. 2B). Taken together, these data suggest that although SARS-CoV-2 infection can induce comparable T cell responses to various viral antigens in young and elderly individuals, pre-existing memory T responses specific to SARS-CoV-2 S and N antigens decrease in elderly people, while M-specific T cell responses are maintained.

**Fig. 2.**
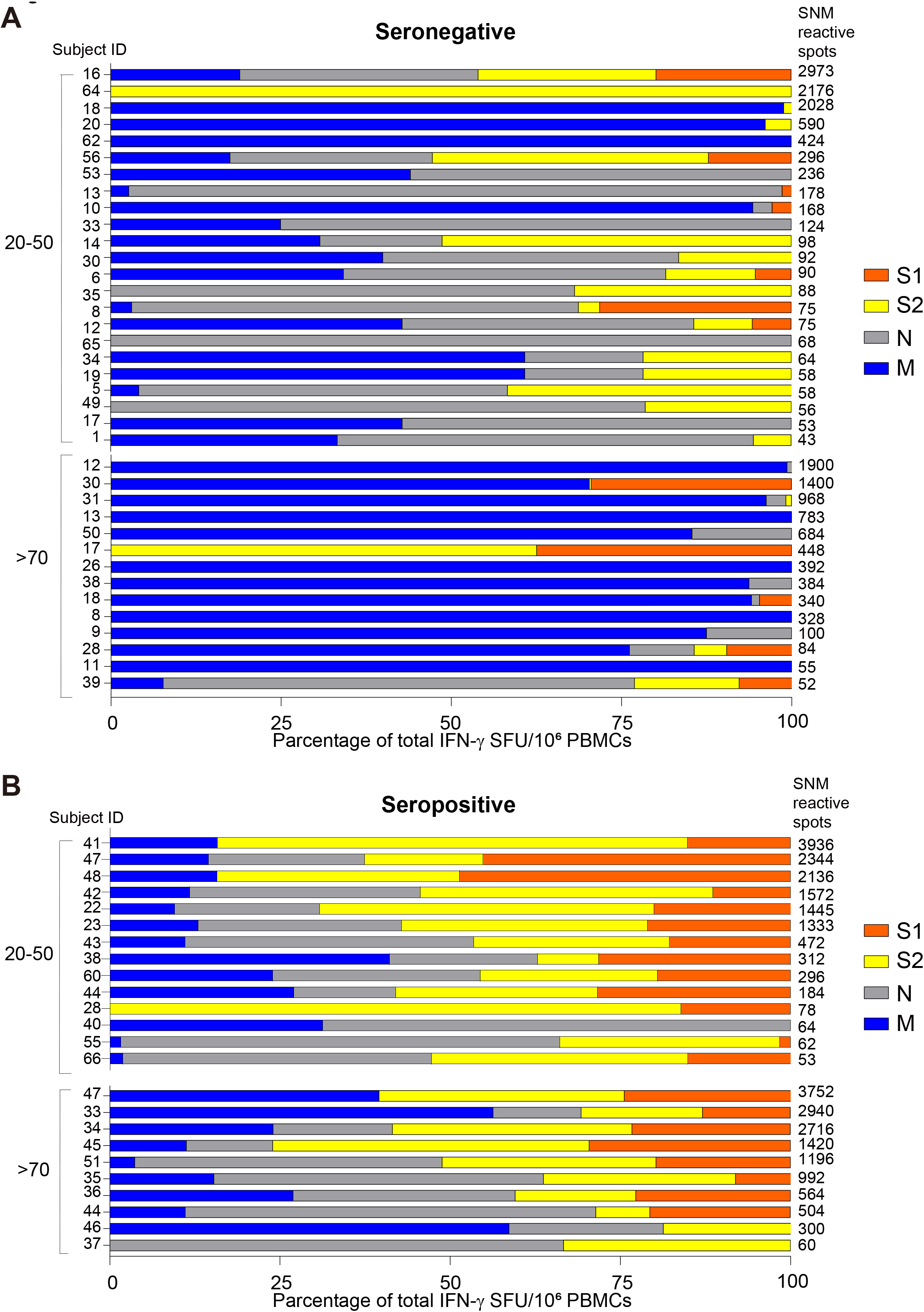
M-specific pre-existing T cell responses predominate in elderly donors. Ratios of spots formed by cells stimulated with SARS-CoV-2 S, N, and M peptide pools in ELISpot data (in Fig. 1.) were analyzed in seronegative (**A**) and seropositive (**B**) donors who had >40 spots/10^6^ PBMCs in the sum of spots formed by cells stimulated with S, N, and M.

### Phenotypes of M-reactive T cells

Given that a proportion of seronegative donors exhibited M-specific responses comparable to those of seropositive donors, we compared phenotypes of M-reactive T cells by flow cytometry analysis. We analyzed 5 seronegative (1 young and 4 elderly) and 5 seropositive (2 young and 3 elderly) donors who had high responses to the M peptide pool (M responders). We stimulated PBMCs with the M peptide pool and stained cells with anti-CD4 or anti-CD8 antibodies, followed by intracellular cytokine staining with anti-IFN-γ antibody. Upon stimulation with the M peptide pool, CD4 T cells but not CD8 T cells significantly increased expression of IFN-γ in both seronegative and seropositive M responders and at comparable levels (Fig. 3A, 3B).

**Fig. 3.**
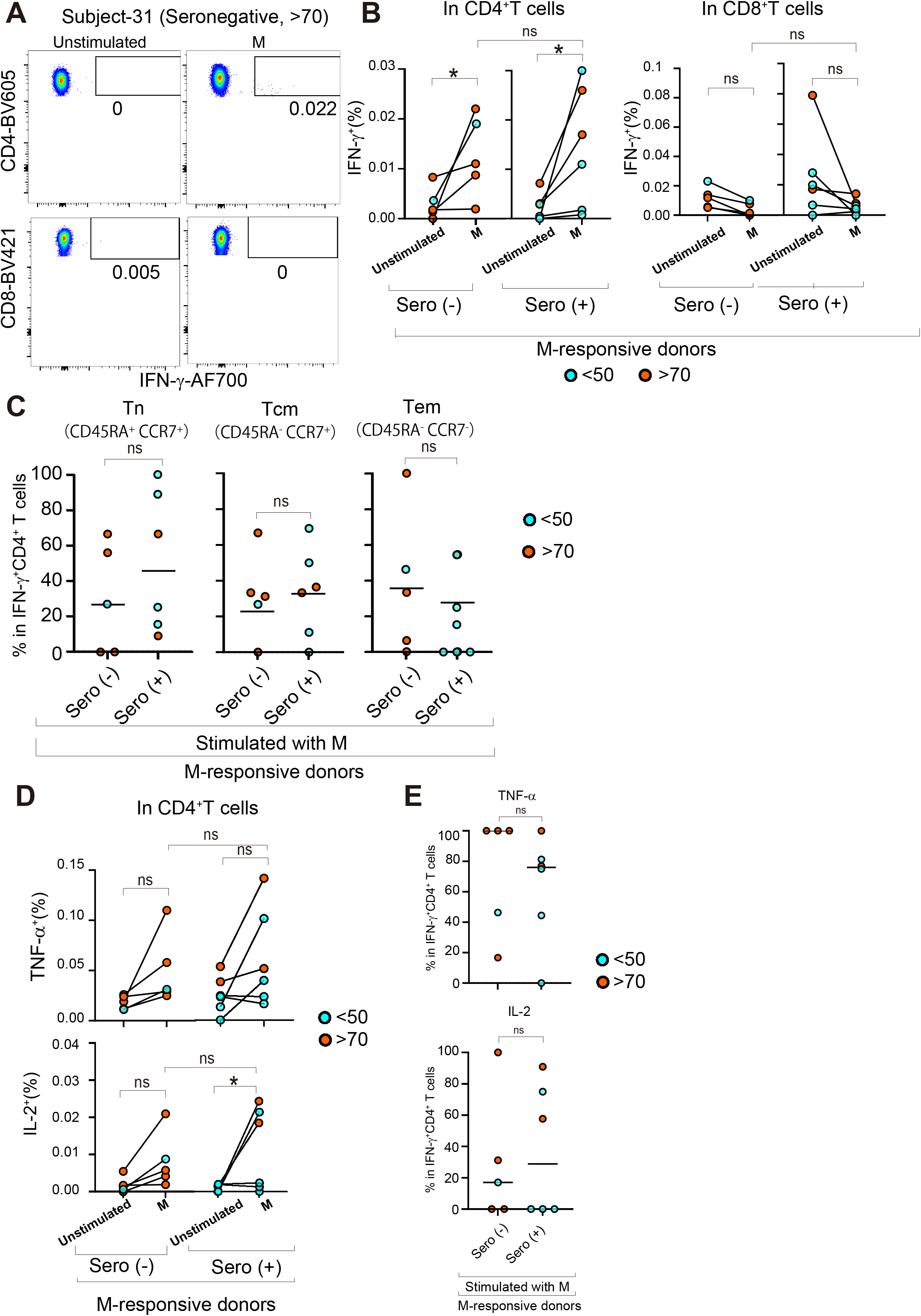
Pre-existing and SARS-CoV-2-induced M-specific T cells exhibit similar phenotypes. PBMCs isolated from seronegative (n=5) and seropositive (n=5) M responders were stimulated with an M peptide pool for 7 hours and analyzed by flow cytometry. The flow cytometry gating strategy was shown in Supplemental Figure 1. (**A**) Dot plots represent IFN-γ expression in CD4 and CD8 T cells of subject #31 (seronegative, >70 years of age). (**B**) Percentages of IFN-γ-expressing cells among CD4 and CD8 T cells were analyzed. (**C**) Percentages of naïve (Tn), central memory (Tcm), and effector memory (Tem) among IFN-γ-expressing cells stimulated with M. (**D, E**) Percentages of TNF-α- and IL-2-expressing cells among total CD4 T cells (D) and IFN-γ-expressing CD4 T cells (E) stimulated with M. (B-E) Data shown as mean ± SD. Each dot represents an individual donor. Blue and orange dots indicate results of young and elderly donors, respectively. Statistical analysis utilized unpaired two-tailed Student’s t tests. *P<0.05, ns: not significant.

We next analyzed CD45RA and CCR7 expression to determine proportions of naïve and memory cells among M-reactive CD4 T cells. Although there was significant variation between individuals in proportions of M-reactive CD4 T cells exhibiting naïve (CD45RA^-^ CCR7^+^), effector memory (CD45RA^+^ CCR7^-^), and central memory (CD45RA^+^ CCR7^+^) phenotypes, all M responders had M-reactive memory T cells (Fig. 3C). There was no obvious difference in the proportion of naïve, effector memory, and central memory CD4 T cells between seronegative and seropositive M responders (Fig. 3C).

We also analyzed frequencies of cells expressing IL-2 and TNF-α among M-reactive CD4 T cells. Although several donors showed increased IL-2 and TNF-α expression upon stimulation with M in both seronegative and seropositive M responders, in seropositive donors, only IL-2 reached statistical significance (Fig. 3D). Seronegative and seropositive M-responders showed no detectable difference in IL-2 and TFN-α expression in T cells stimulated with M (Fig. 3D). Expression of TNF-α and IL-2 in IFN-γ-expressing CD4 T cells was comparable between seronegative and seropositive M responders (Fig. 3E). These data indicate that M-reactive T cell responses are mediated by CD4 T cells expressing IFN-γ in both seronegative and seropositive M responders, suggesting that pre-existing M-reactive T cells and SARS-CoV-2-induced memory M-specific T cells might serve similar functions in SARS-CoV-2 infection.

### Epitopes of SARS-CoV-2 M protein

To compare epitopes recognized by M-reactive T cells of seronegative and seropositive donors, we stimulated PBMCs isolated from M responders with SARS-CoV-2 M matrix pools (15 pools of 6-8 peptides) where each of 56 M-derived peptides (15-mers) is allocated to 2 different pools. In seropositive M responders, stimulation with pools 5 and 13 induced high levels of IFN-γ responses (Fig. 4A). These matrix pools shared a single peptide, M_145-160_, suggesting that the M_145-160_, which was previously identified as an immunodominant viral epitope in COVID-19 convalescent patents (18), is the epitope recognized by T cells of seropositive M responders. In contrast, in seronegative M responders, stimulation of PBMCs with many M matrix pools induced comparable IFN-γ responses (Fig. 4B), suggesting that various M epitopes are recognized by pre-existing T cells.

**Fig. 4.**
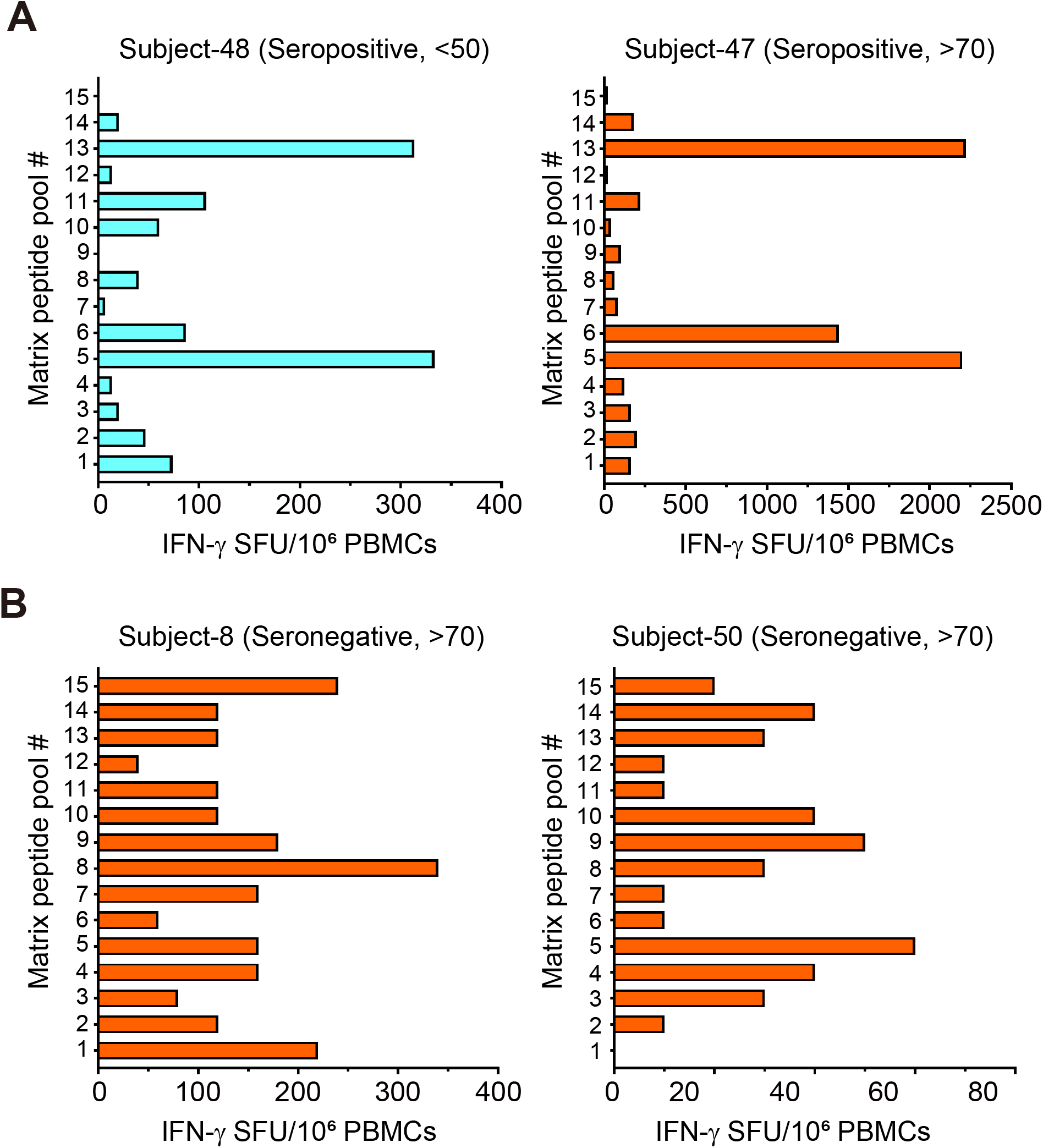
SARS-CoV-2 M epitopes recognized by T cells. PBMCs isolated from M responders in seropositive (**A**) and seronegative (**B**) groups were stimulated with SARS-CoV-2 M matrix pools (15 pools) for 16 h and subjected to IFN-γ ELISpot analysis. Spot-forming units representing the frequency of IFN-γ-secreting cells. Blue and orange bars indicate results of young and elderly donors, respectively.

## Discussion

Our data indicate that a fraction of elderly donors possess significantly high levels of pre-existing SARS-CoV-2 M-specific T cell responses, though the frequency of pre-existing SARS-CoV-2 S- and N-specific T cells in this population is lower. Other recent studies have also reported an age-related decline of S-specific pre-existing T cells (Refs. 19 and L. Loyal et al., manuscript posted on medRxiv DOI: 10.1101/2021.04.01.21252379). These data suggest that pre-existing T cells specific to SARS-CoV-2 are heterogeneously affected by age in a target antigen-dependent manner. There was no obvious difference in the sum of IFN-γ-expressing cells specific to S, N, and M antigens, suggesting that abundant M-specific T cells can compensate for the loss of S- and N-specific T cells, at least in the magnitude of T cell-mediated IFN-γ production, in elderly individuals. Taken together, these data suggest that the diversity of target antigen repertoire for pre-existing T cells may decline with age, but the magnitude of pre-existing T cell responses can be maintained with T cells specific to certain viral proteins such as M.

As the frequency of pre-existing T cells specific to viral structural proteins S, N, and M is associated with protection from SARS-CoV-2 infection (D. Wyllie et al., manuscript posted on medRxiv DOI: 10.1101/2020.11.02.20222778), focused M responses might be particularly important for protection of elderly individuals who have lower responses to S and N. Flow cytometry analyses revealed that CD4 T cells mainly mediate M-specific T cell responses, and their naïve/memory phenotypes and their capacity to produce IFN-γ, IL-2, and TNF-α cytokines were comparable between seronegative and seropositive groups. The phenotypic similarity suggests that pre-existing M-reactive T cells may serve similar functions to SARS-CoV-2-induced M-specific memory T cells. We speculate that pre-existing M-specific CD4 T cells play a protective role in SARS-CoV-2 infection by promoting cellular immunity through IFN-γ production and humoral immunity by providing T cell help to S- and N-specific B cells via linked recognition.

However, we cannot exclude the possibility that pre-existing M-specific T cells are harmful for some elderly individuals. Several studies suggest that pre-existing SARS-CoV-2-specific T cells are detrimental in COVID-19 (8). In particular, the frequency of M-specific T cells in COVID-19 patients is thought to be a risk factor, as it is correlated with age and severity of disease (8), although how pre-existing M-specific T cells affect magnitude, kinetics and functions of M-specific T cell responses in COVID-19 patients remains unclear. Thus, both protective and pathogenic functions of pre-existing M-specific T cells can be speculated. A longitudinal comparison of susceptibility and symptom severity of COVID-19 between individuals with and without high pre-existing M-specific T cell responses might provide insights into this issue.

Despite defects in pre-existing T cell responses to S and N, most elderly donors who recovered from mild COVID-19 had abundant T cells specific to S and N antigens at levels comparable to those of young donors, suggesting that elderly individuals can induce T cell responses against S and N antigens upon SARS-CoV-2 infection. However, our analysis is limited to only a few patients who had recovered from mild COVID-19. Therefore, the relationship between age-related alteration of pre-existing T cells and T cell responses during infection in patients with diverse clinical outcomes of COVID-19 should be investigated in a larger, statistically valid test population.

Whether diverse SARS-CoV-2-induced T cell clones can mediate long-lasting memory responses should be addressed in future studies. It has recently been shown that SARS-CoV-2-induced memory T cells persist at least 6 months after infection (20). Interestingly, SARS-CoV-1 infection induces long-lasting (>11 years) CD8 memory T cells specific to M_141-155_ peptide (21). Our data show that the overlapping peptide (M_145-160_) is immunodominant in SARS-CoV-2 infection, which is consistent with a recent study showing CD4 T cell responses to the M_145-160_ peptide in convalescent COVID-19 patients (18). The amino acid sequence of M_145-160_ peptide from SARS-CoV-2 shows high homology with SARS-CoV-1 and other coronaviruses (SARS-CoV-1: 81.3%, NL63: 33.0%, OC43: 47.0%, 229E: 22.7%, HKU1: 47.0%). This short M peptide is likely a potent inducer of SARS-CoV-2 M-specific memory T cells. In contrast, pre-existing T cells likely recognize various M peptides, possibly including M_145-160_, rather than focusing on this single M peptide, as we observed in epitope mapping analysis.

What induces pre-existing M-specific T cells? Common cold coronaviruses may induce pre-existing SARS-CoV-2-specific T cell (22, 23). Amino acid sequence homology between SARS-CoV-2 and other common cold coronaviruses is relatively high for M (NL63: 25.2%, OC43: 36.9%, 229E: 26.7%, HKU1: 32.4%), S1 (NL63: 11.0%, OC43: 15.4%, 229E: 12.8%, HKU1: 15.2%), S2 (NL63: 27.3%, OC43: 36.9%, 229E: 28.0%, HKU1: 35.3%), and N (NL63: 22.3%, OC43: 26.5%, 229E: 16.2%, HKU1: 26.5%). These coronaviruses may induce polyclonal M-specific T cells. Age-related loss of memory T cells specific to common cold coronavirus S protein (19) supports the hypothesis that pre-existing M-focused T cell responses are induced by common cold coronavirus infection in elderly people. Our data showing higher frequency of pre-existing T cells specific to S-2 than S-1 are also consistent with the fact that S-2 shows higher homology between SARS-CoV-2 and other coronaviruses. However, some young donors, as well as elderly donors, had abundant pre-existing T cells specific to M, but not to S and N, suggesting that focused T cell responses to M are not necessarily due to age-related loss of pre-existing T cells specific for S and N antigens. Interestingly, a recent study reported that T cells specific to commensal bacteria can cross-react with SARS-CoV-2 S antigen (X. Lu et al., manuscript posted on medRxiv DOI: 10.1101/2021.03.23.436573). Similarly, there may be specific microbes that induce pre-existing M-specific T cells.

It is worth considering the potential of novel COVID-19 vaccines to induce M-specific immunity. Current vaccine strategies are to induce S-specific antibody and T cell responses (24, 25). Recent studies reported a correlation between the frequency of pre-existing S-specific T cells and vaccine-induced S-specific T cell responses (L. Loyal et al., manuscript posted on medRxiv DOI: 10.1101/2021.04.01.21252379), which suggests a role of pre-existing S-specific T cells in cognate T cell help. However, elderly individuals likely would not benefit fully from pre-existing S-specific T cells. To enhance vaccine efficacy among the elderly, it might be reasonable to consider a strategy to induce not only S-specific, but also M-specific immunity, using vaccines based on inactivated viruses or M-fused S antigens. Linked recognition of M-specific T helper cells by S-specific B cells can promote S-specific antibody production by overcoming the defect of cognate T cell help in elderly individuals. Further characterization of M-specific T cells in young and elderly may provide new insights into vaccine-induced immunity that is less affected by age.

## Data Availability

our data do not have availability links

## Acknowledgements

We thank physicians and nurses at KIN oncology Clinic for excellent support to collect blood samples from donors. We also thank Steven Aird for editing manuscript. We are also grateful to OIST Graduate University for its generous funding of the Immune Signal Unit.

## Notes

### Competing Interest Statement

The authors have declared no competing interest.

### Funding Statement

OIST Graduate University for funding of the Immune Signal Unit.

### Author Declarations

Okinawa Institute of Science and Technology, Graduate University (OIST) human subjects ethics committee (applications HSR-2020-024, HSR-2020-028)

