## supplemental Figure 1 for "Altered pre-existing SARS-CoV-2-specific T cell responses in elderly individuals"

Suplemental Figure 1

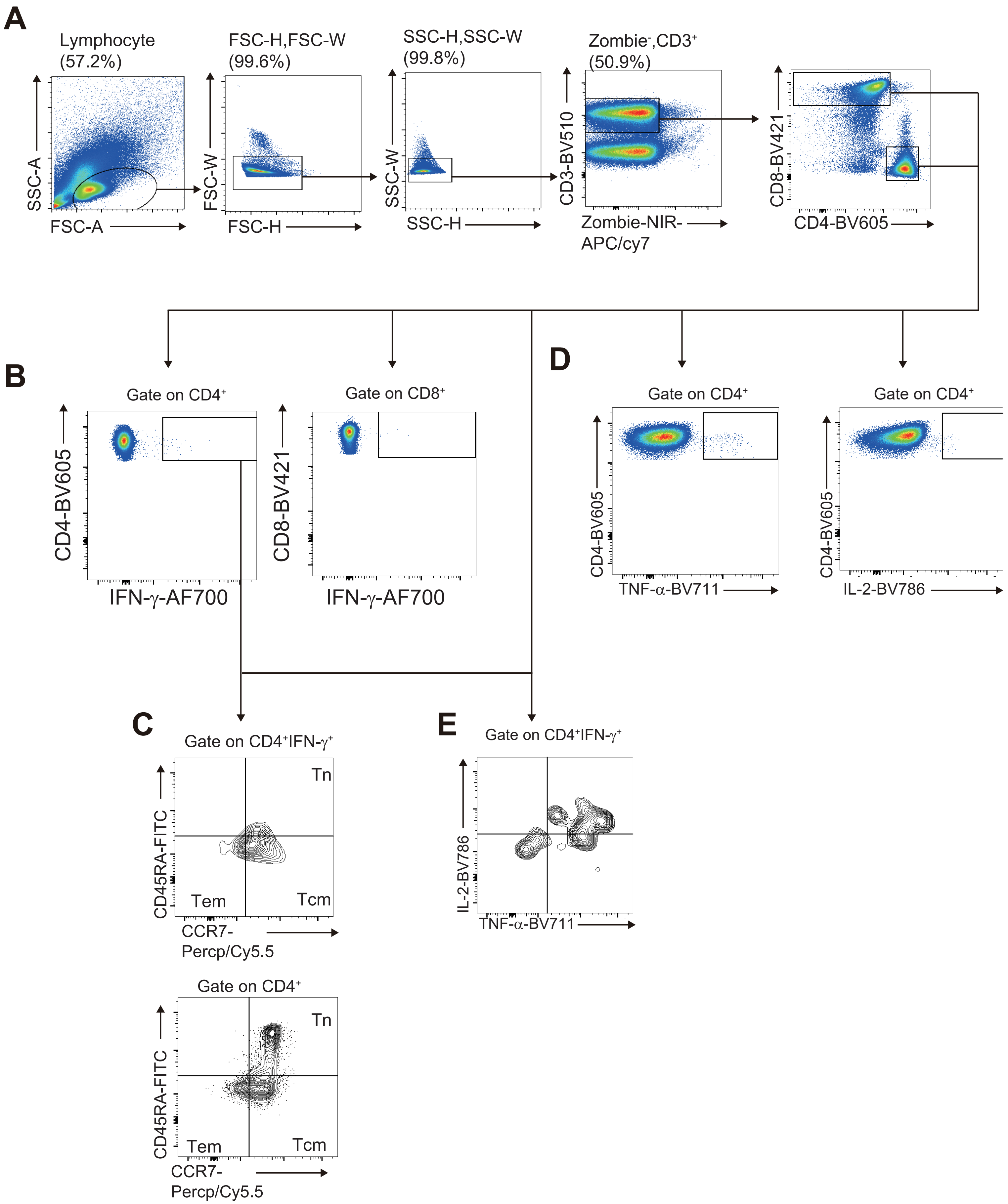


**Supplemental Figure 1. Gating strategy, Related to Figure 3**

Flow cytometry gating strategy for living (NIR-Zombie$^-$) CD4$^+$ and CD8$^+$ T cells gated on CD3$^+$ (A), IFN-γ-expressing cells gated on CD3$^+$CD4$^+$ or CD3$^+$CD8$^+$ T cells (B), naïve (Tn: CD45RA$^+$CCR7$^+$), central memory (Tcm: CD45RA$^-$CCR7$^+$) and effector memory (Tem: CD45RA$^-$CCR7$^-$) T cells gated on CD3$^+$CD4$^+$ or CD3$^+$CD4$^+$ IFN-γ$^+$ (C), TNF-α- and IL-2-expressing T cells gated on CD3$^+$CD4$^+$ (D) or CD3$^+$CD4$^+$IFN-γ$^+$ (E).
